# Stress from Cadaver Dissection Linked to Learning Conditions: Evidence from Sub-Saharan Africa

**DOI:** 10.1101/2024.07.09.24310166

**Authors:** Jude Amechi Nnaka, Victor Udochukwu Ezeike, Kristjan Thompson, Izuchukwu Azuka Okafor

**Affiliations:** Department of Anatomy, Faculty of Basic Medical Sciences, College of Health Sciences, Nnamdi Azikiwe University, Nnewi Campus, PMB 5001, Nnewi, Anambra State, Nigeria; Department of Mental Health, University of Abuja Teaching Hospital, Gwagwalada-Zuba, 902101, Gwagwalada, P.M.B. 228, Abuja, Federal Capital Territory, Nigeria; Department of Biomedical Sciences, Mercer University School of Medicine, Savannah, GA, USA

**Author notes:** **Corresponding author**: Dr. Izuchukwu Azuka Okafor, Department of Anatomy, Faculty of Basic Medical Sciences, College of Health Sciences, Nnamdi Azikiwe University, Nnewi Campus, PMB 5001, Nnewi, Nigeria.

**Keywords:** anatomy education, cadaver dissection, learning conditions, stress, teaching method

## Abstract

**Objective:** This study examined the relationship between students’ perceptions of cadaver dissection (CD) and some learning conditions during CD.

**Methods:** This is a cross-sectional study involving 2968 medical students and graduates exposed to CD at nine countries systematically and proportionally selected from Sub-Saharan African countries based on their ranking on the four World Bank indices of development and education: population, literacy, human capital index (HCI), and human development index (HDI). A self-administered questionnaire was used to collect relevant data on learning conditions during CD experience of the participants, using online channels, including email, Whatsapp, Facebook, Instagram, and Twitter.

**Results:** About half (48.76%) of the participants perceived CD as stressful, while 51.24% thought otherwise. However, 57% of participants from institutions where a CD session lasts three hours described their experience as stressful, whereas 69.67% of participants whose institution spent one hour in a single CD session agreed that CD is not stressful. Similarly, 60.63% of participants from institutions with a student-donor ratio between 5 to 10 students per cadaver described their experience as ‘not stressful’. In comparison, 57.51% of participants from institutions with a student-donor ratio of 10–20 students per donor and 53.80% of participants from institutions with over 20 students per donor described their experience as stressful.

**Conclusions:** Students from institutions with CD instructors, shorter CD sessions, and a smaller student-donor ratio are more likely to perceive CD as stress-free. Providing more support for students during CD and reducing time for CD may be an antidote to CD-related stress.

## Introduction

Cadaver dissection (CD) is an age-long anatomy-teaching method that has stood the test of time.^1–2^ Since its first use as a pedagogical tool in the thirteenth century^3^, it has undergone modifications such as the introduction of prosections and plastination^4–5^, with recent technology-driven integration like digital imaging technology, virtual dissection, and 3D printing.^6–7^ While these new methods offer advantages such as accessibility, convenience, and the ability to learn without the need for a donor^8–9^, CD is still considered by many modern scholars and educators to be an indispensable tool in medical education, as it offers a thorough understanding of the human body’s complexities through an immersive and practical learning experience that is impossible to achieve through virtual simulations alone.^10–11^

CD, despite its benefits and importance in anatomy education, is not without its unique challenges. Among these challenges is the issue of anxiety, mental stress, and emotional distress^12–14^ which is considered to be a side effect of working closely with deceased human bodies.^14–15^ These challenges are often compounded by the pressure to pass exams and very high academic workloads. Additional stressors due to the scarcity of donors have been shown to contribute to these challenges, particularly in low-income countries, where there may be reduced opportunities for every student to have ample hands-on experience during CD.^14,16^

Medical and allied health science students encounter numerous stressors throughout their training, including those associated with CD. While many students can navigate CD-related stressors effectively, a subset may find the experience highly challenging and overwhelming.^15,17^ These variations in students’ responses to CD-related stress are influenced by diverse factors, including past traumatic experiences, prior medical exposure, and individual resilience levels.^12,17–20^ Studies targeting the reduction and management of CD-related stress have emerged, with authors presenting various recommendations such as providing humanizing information about the donor^1^, carrying out desensitizing pre-CD sessions^19,21–23^ and the use of background music during CD.^24^

Considering these known strategies, questions persist on the impact of specific learning conditions on CD-related stress and management. While previous studies have found that the perception of one’s learning environment is associated with the academic performance and well-being of medical students^25^, no study has specifically examined the relationship between learning conditions during CD and students’ stress levels. Understanding this relationship is crucial to improving student’s CD experience. Medical educators can implement strategies to mitigate these stressors and create a more supportive learning environment by identifying the factors that contribute to elevated stress levels during CD. This study investigated the poor learning conditions during CD and examines their relationship with students’ perceptions of CD.

## Methods

### Study plan, participant recruitment, and data collection

This cross-sectional study assessed the relationship between CD learning conditions and students’ perception of CD. The study systematically and proportionally targeted students and graduates from Sub-Saharan African countries based on their ranking on the four World Bank indices of development and education - population, literacy, human capital index (HCI), and human development index (HDI) - to avoid selection bias and to ensure that the countries that have more medical schools, CD-based programs, students, and medical graduates are reached. The country scoring and selection based on the above rankings have been earlier published.^26^ After ranking, the countries that scored the highest were selected for inclusion till 20% of countries were included, with at least one country from each Sub-Saharan region. A total of 9 countries were selected – Cameroon, Ghana, Kenya Malawi, Nigeria, South Africa, Uganda, Zambia, and Zimbabwe - with a total of 2968 respondents with a sample size ranging from 150 to 788 in each country. The data for all the countries has been published in an open repository.^26^ Only students or graduates who had dissection experience in any institution within the selected countries were included in this study. Students without CD experience were excluded to reduce bias and ensure reliability. This study considered a wide range of students and graduates from different medical science and health fields who completed an online self-administered questionnaire shared through different platforms (e-mails, texts, WhatsApp, Facebook, and Twitter).

### Study questionnaire and variables

This study utilized an online self-administered questionnaire (https://forms.app/judennaka/BM-in-dissection), with questions focusing on the key objectives of the study. A pilot survey was done with a heterogeneous population of 15 participants, and modifications were made to the questionnaire following the outcome. The final questionnaire draft was validated using internal consistency (Cronbach’s alpha of 0.7), test-retest reliability, and inter-rater reliability (Kappa statistic, K = 0.9) before commencement of data collection.

### Statistical Analysis

The study data were automatically generated from the site where the questionnaire was hosted and were prechecked for errors using Openrefine version 3.6 before transfer to IBM SPSS version 25 for analysis. Descriptive analysis, Chi-squared test, and one-sample non-parametric test were used to determine proportions, association, and one-sample differences respectively. Data analysis was conducted with significance set at p<0.05.

## Results

### Stress Perception during Cadaver Dissection

The perception of stress during CD remains uncertain, as 51% of respondents assert that it is not stressful, while 48.36% agree that it induces stress (Figure 1).

**Figure 1:**
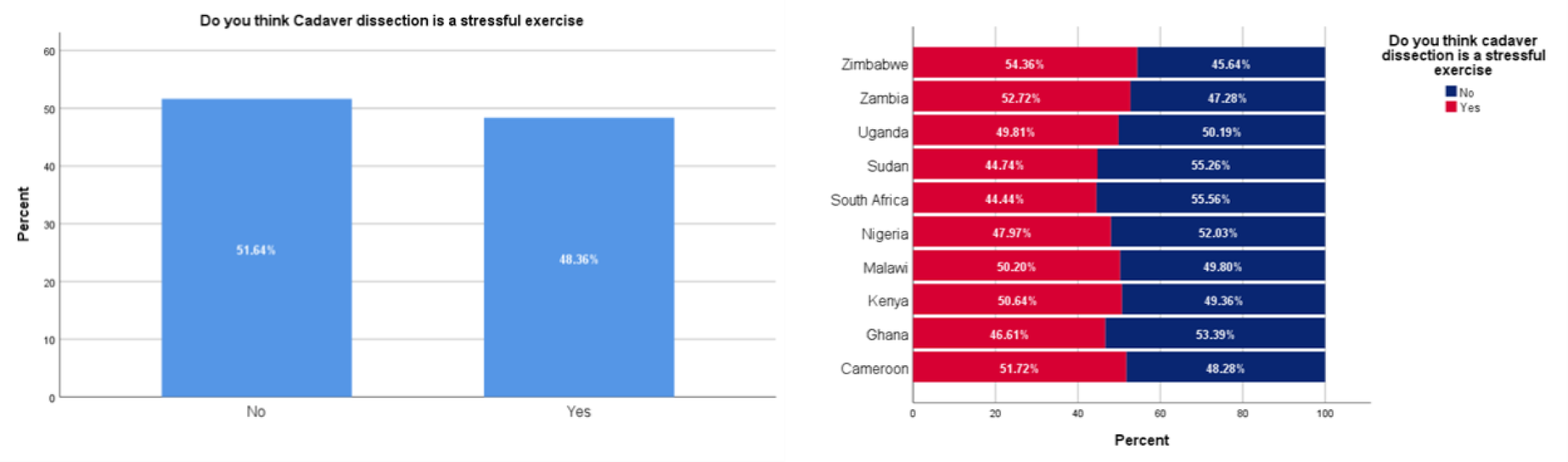
Student’s stress perception during CD.

### Cadaver Dissection Learning Conditions

The duration of CD sessions in most institutions is predominantly between 2 hours (46.77%) and 3 hours (29.41%), with less than 2% of institutions reporting CD sessions lasting less than 1 hour (Figure 2). Student-donor ratios vary, with 39.12% having 5-10 students, and 29.57% having 10-20 students per donor. About 21% of students reported a ratio exceeding 20 students per donor, while only 8.13% reported a ratio of less than five students per donor (Figure 3).

**Figure 2:**
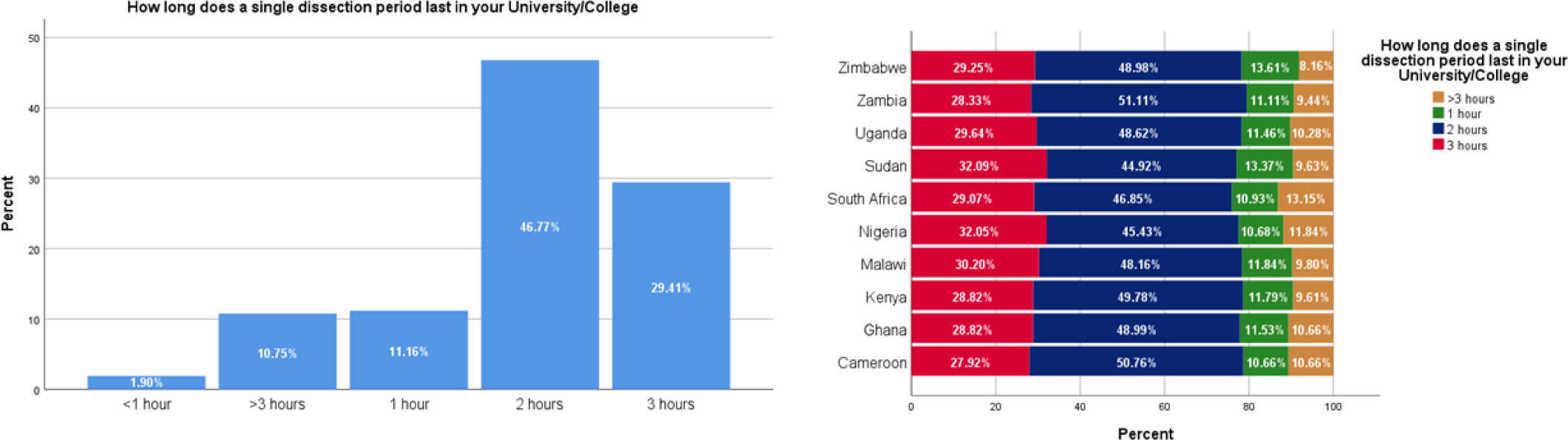
CD Duration in sub-Saharan African medical schools.

**Figure 3:**
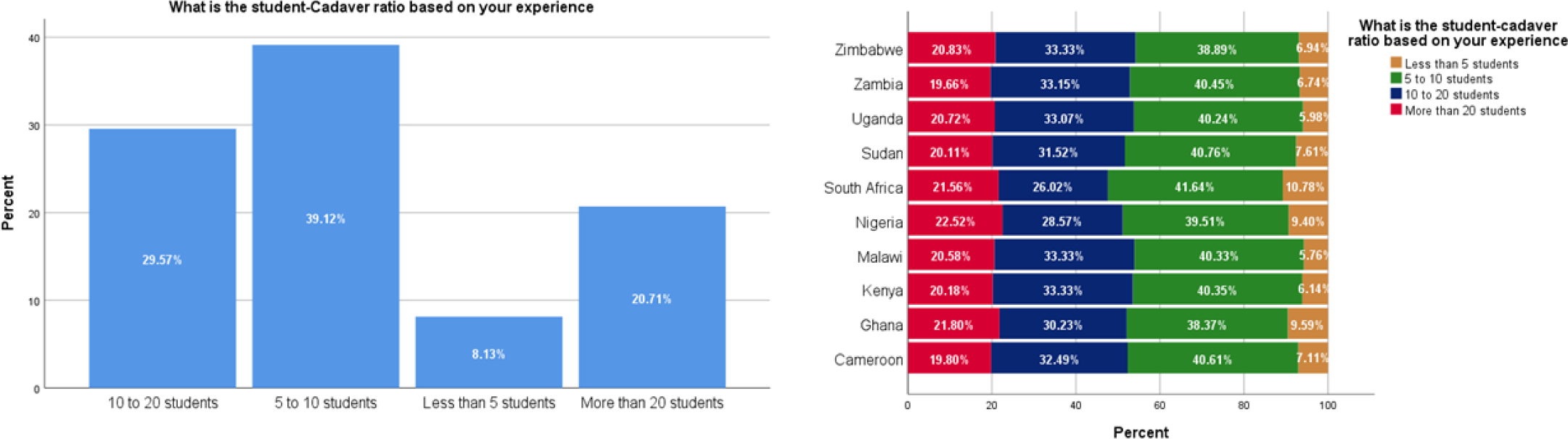
Student-donor ratio in sub-Saharan African medical schools.

Regarding supervision, 35.71% of respondents reported always having tutors present during CD, while 24.29% and 24.43% reported having tutors often and sometimes, respectively. Less than 2% reported never having a tutor during CD sessions (Figure 5).

**Figure 4:**
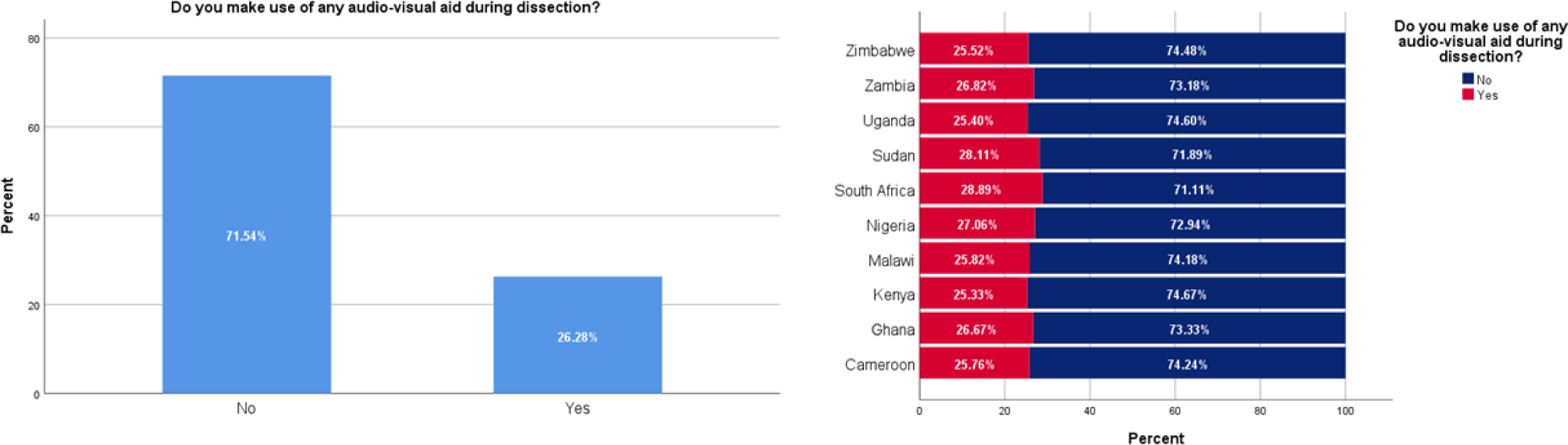
Prevalence of the use of audio-visual aids during CD in sub-Saharan medical schools.

**Figure 5:**
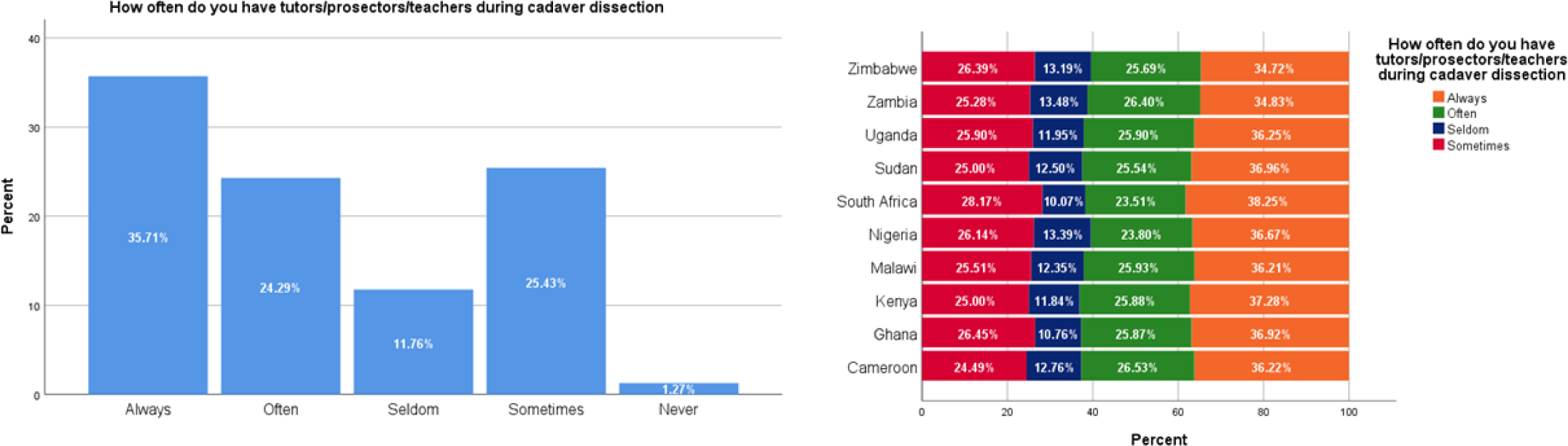
Availability of tutors/instructors during CD in sub-Saharan African medical schools.

Audiovisual aids were not utilized in the majority (71%) of institutions, with only 26.26% confirming their use during CD (Figure 4). In terms of safety measures, 66.86% of respondents reported consistently using personal protective equipment (PPE) during dissection, while 9.20% admitted to never using PPE during the process (Figure 6).

**Figure 6:**
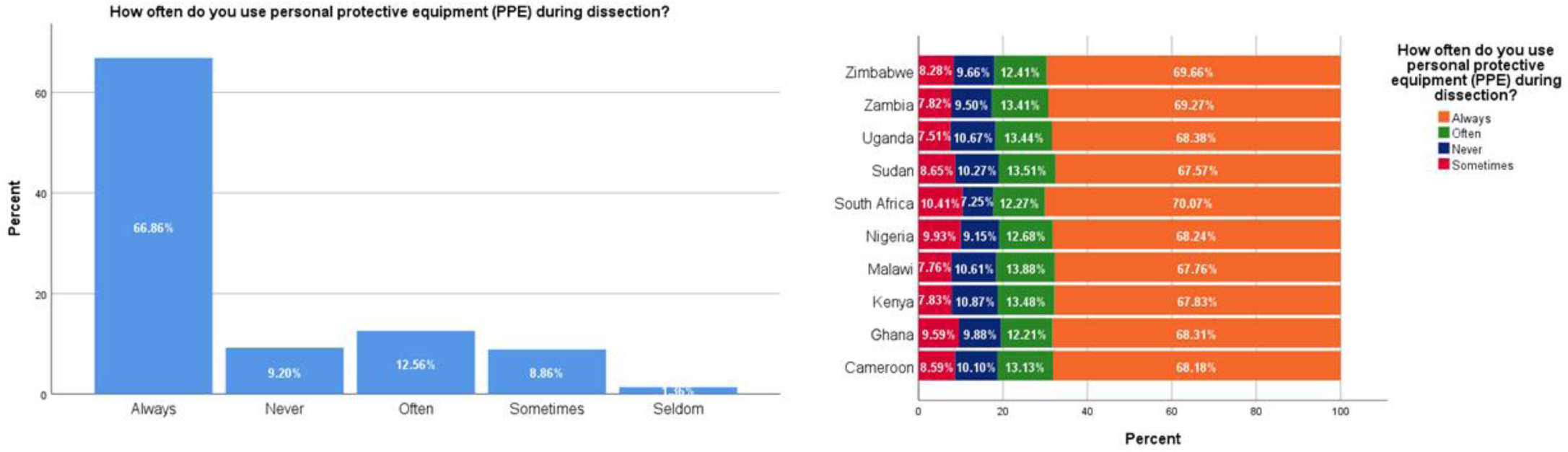
The Use of PPEs by students during CD in sub-Saharan African medical schools.

### Relationship Between Learning Conditions and Stress Perception in Cadaver Dissection

There was also a significant association (p<0.001) between student-donor ratio and stress perception in CD. Respondents in CD sessions with a student-donor ratio of less than 5 students predominantly perceived CD as not stressful, while those with ratios of 10 to 20 students and more than 20 students showed a higher proportion perceiving it as stressful.

The duration of CD sessions showed a statistically significant relationship with the participants’ stress perception (p<0.001). Sessions lasting less than 1 hour were predominantly associated with low-stress responses, whereas sessions exceeding 3 hours had a higher proportion of respondents perceiving CD as stressful (p<0.001).

Stress perception in CD exhibited a significant association (p<0.001) with the presence or frequency of tutors. Participants who reported always having tutors during CD were more likely to perceive it as not stressful, whereas those who reported “seldom” or “sometimes” having tutors showed a higher proportion of perceiving it as stressful. The association between stress perception of CD and the utilization of audio-visual aids was found to be significant (p<0.001). Respondents using audio-visual aids during CD were more likely to perceive it as not stressful, while those who did not use such aids showed a higher proportion of perceiving CD as stressful.

The association between stress perception in CD and the frequency of personal protective equipment (PPE) use was significant (p<0.001). Participants reporting always using PPE during dissection were more likely to perceive it as not stressful, while those reporting never or sometimes using PPE showed a higher proportion of perceiving it as stressful.

## Discussion

This study explores students’ perceptions of stress during CD and some of the learning factors that could contribute to their stress levels. The result from this study shows that students’ views on CD were evenly split between perceiving it as stressful or not stressful. This suggests that stress levels during CD are likely subjective and influenced by both internal and external factors (citation needed). This finding is consistent with prior research, which suggests that responses to stress during CD are influenced by factors such as gender, previous traumatic experiences, and prior medical experiences.^12,17^

Most of the surveyed students reported that CD sessions last between 2 to 3 hours, a duration comparable to findings from studies conducted in Australia^27^ and South Korea.^28^ This suggests a commonality in the timeframe of CD sessions across different geographical regions, indicating standardized approaches to teaching gross anatomy in Sub-Saharan Africa. However, student-donor ratios in this presented study are high, with nearly 30% having 10-20 students per donor and 21% of students reporting a concerning ratio exceeding 20 students per donor. This distribution is significantly higher than the student-donor ratio of 5 students per donor reported in US and Canadian institutions.^29–31^ This reflects the challenges faced by institutions in resource-constrained settings, where large class sizes may be necessitated by limited donor availability and funding constraints. This issue is further compounded by the very low utilization of audiovisual aids during CD in sub-Saharan African institutions in this study (less than 2%). These disparities in access to technology and educational resources, which are more pronounced in Sub-Saharan Africa compared to more economically developed regions^31^, could hinder the enhancement of learning experiences and the development of a visual understanding of anatomical structures among students.

In examining the relationship between learning conditions and students’ perceptions of stress during CD, this study found a significant correlation between the duration of CD sessions and students’ stress perception. Specifically, nearly 70% students who experienced CD sessions lasting for only one hour perceived it positively (Table 1). On the other hand, 57% of participants from institutions where a CD session lasts three hours described their experience as stressful. This suggests that there may be a threshold for how long students can effectively engage with CD before it becomes overwhelming. Previous studies have shown that not only do long working hours increase the likelihood of occupational stress, but they also have a direct positive effect on mental disturbances like depression.^32–33^ Longer CD sessions, combined with the emotional and psychological challenges they present, may contribute to physical and mental fatigue, resulting in increased actual and perceived stress levels among students.^34–35^ It could be beneficial for educators to consider breaking up CD sessions into shorter intervals to help students manage their stress levels and maintain focus.

**Table 1:** The relationship Between Learning Conditions and Stress Perception in Cadaver Dissection.

|  |  | Do you think<br>Cadaver dissection<br>is a stressful<br>exercise |  | Total | P value |
| --- | --- | --- | --- | --- | --- |
|  |  | No | Yes |  |  |
| What is the student-donor<br>ratio based on your<br>experience? | 10 to 20 students | 397 | 538 | 935 |  |
|  | 5 to 10 students | 750 | 487 | 1237 | P< 0.001* |
|  | <5 students | 146 | 111 | 257 |  |
|  | >20 students | 303 | 352 | 655 |  |
|  | Total | 1596 | 1488 | 3084 |  |
| How long does a single dissection period last in your University/College? | <1 hour | 39 | 21 | 60 |  |
|  | >3 hours | 168 | 172 | 340 |  |
|  | 1 hour | 246 | 107 | 353 | P< 0.001* |
|  | 2 hours | 780 | 699 | 1479 |  |
|  | 3 hours | 400 | 530 | 930 |  |
|  | Total | 1633 | 1529 | 3162 |  |
| How often do you have tutors/prosectors/teachers during cadaver dissection? | Always | 681 | 448 | 1129 |  |
|  | Never | 21 | 19 | 40 | P< 0.001* |
|  | Often | 391 | 377 | 768 |  |
|  | Seldom | 141 | 231 | 372 |  |
|  | Sometimes | 381 | 423 | 804 |  |
|  | Total | 1615 | 1498 | 3113 |  |
| Do you make use of any audio-visual aid during dissection? | No | 1103 | 1159 | 2262 | P< 0.001* |
|  | Yes | 503 | 328 | 831 |  |
|  | Total | 1606 | 1487 | 3093 |  |
| How often do you use personal protective equipment (PPE) during dissection? | Always | 1154 | 960 | 2114 |  |
|  | Never | 137 | 154 | 291 | P< 0.001* |
|  | Often | 170 | 227 | 397 |  |
|  | Seldom | 25 | 18 | 43 |  |
|  | Sometimes | 137 | 143 | 280 |  |
|  | Total | 1623 | 1502 | 3125 |  |

Data was analyzed using student's Chi-squared test and values are considered significant at
$P < 0.05^*$

The level of guidance and support offered during CD sessions significantly influences student’s perceptions of CD. Those who reported receiving more guidance and support from instructors were less likely to perceive CD as stressful. This highlights the crucial role of active and engaged educators during CD to support students. A study by Manyama and colleagues^36^ found that engaged and reciprocal teaching in CD not only improves students’ performance but also boosts their confidence and helps them to communicate better. Also, Trigueros and coworkers^37^ demonstrated that encouraging autonomy and providing support for students significantly reduces academic stress. These findings suggest that the role of educators in CD goes beyond simply teaching anatomy, but also includes providing emotional support and creating a supportive learning environment for students.

Access to visual aids and videos during CD also reduces the likelihood of perceiving CD as stressful. It is known that visual aids and videos provide a more comprehensive understanding of the CD process, allowing students to visualize and better comprehend the task at hand.^38^ Also, the visual aids and videos may serve as a source of reassurance and guidance, helping to alleviate any anxiety or uncertainty that students may experience during CD.^39^

Results from this study show a significant relationship between class size and stress levels during CD. Students in classes with 10 - 20 students or more per donor were more likely to perceive CD as stressful compared to students in classes with less than 10 student-donor ratios. This suggests that larger class sizes may contribute to heightened stress levels during CD, potentially due to factors such as increased competition for access to resources, reduced individualized attention from instructors, and a higher likelihood of feeling overwhelmed in larger groups.^36^ While there is yet to be any consensus on the ideal student-donor ratio for CD, it has been shown elsewhere that a team of three to four students per donor is adequate to carry out dissection tasks.^36,40^

Taken together, this study demonstrates that teaching duration, class size, instructional support, and educational resources can shape students’ stress experiences during CD. As such, medical educators must continually review their learning conditions and strategies to ensure that CD-related stress is reduced to the barest minimum. This review could include ensuring that the time allocated for a CD session is within 2 to 3 hours, increasing access to tutors during CD, and reducing to student-donor ratio to not more than 10 per donor. By addressing these elements, students can feel more confident and supported throughout the dissection process, ultimately enhancing their educational experience and overall well-being.^30^

This study provides valuable insights into the relationship between learning conditions and students’ stress perception during CD; however, there exists some limitations. Firstly, the survey relied on self-reporting, which introduces the possibility of response bias and inaccuracies in data collection. Additionally, the cross-sectional design of this study restricts the ability to establish causality or temporal relationships between the study variables. Therefore, it is important to interpret the results of this study with caution and consider the possibility of confounding variables such as individual differences in stress coping strategies or the influence of participant’s previous experiences in CD on the study outcomes. Future research employing longitudinal designs and diverse participant samples could provide a more comprehensive understanding of the factors influencing students’ stress perception during CD.

## Conclusion

This study shows that students from institutions with CD instructors, shorter CD sessions, and a smaller student-donor ratio are more likely to perceive CD as stress-free. This suggests that institutions that prioritize creating a comfortable and inclusive learning environment during CD sessions are more likely to have reduced levels of stress among students. Providing more support for students during CD and reducing time for CD may be an antidote to CD-related stress.

## Source of Funding

This research did not receive any specific grant from funding agencies in the public, commercial, or not-for-profit sectors.

## Conflict of Interest

Authors have no conflict of interest to declare

## Ethical Approval

This study obtained ethical clearance from the Nnamdi Azikiwe University Faculty of Basic Medical Science Ethical Committee with the approval number NAU/CHS/NC/FMBS/663. The study also complied with all the ethical guidelines of Helsinki Declaration and guidelines for questionnaire and surveys of the National Health Research Ethics Committee of Nigeria (NHREC) guidelines.

## Consent

The online survey used for the study was anonymized and was only accessed by the participants upon submitting an informed consent.

## Author Contributions

IAO conceived and designed the study. JAN and IAO collected and analyzed the data. All the authors participated in the data interpretation, writing and revision of the manuscript. All authors have critically reviewed and approved the final draft and are responsible for the content and similarity index of the manuscript.

## Data Availability

All data produced in the present study are available upon reasonable request to the authors

https://figshare.com/articles/dataset/Use_of_background_music_and_cadaver_dissection-associated_stress_in_Sub-Saharan_Africa/24271675

## Notes

### Competing Interest Statement

The authors have declared no competing interest.

### Funding Statement

This study did not receive any funding

### Author Declarations

This study obtained ethical clearance from the Nnamdi Azikiwe University Faculty of Basic Medical Science Ethical Committee with the approval number NAU/CHS/NC/FMBS/663

